# Masticatory Performance and Mortality in Individuals with Cardiovascular Diseases: A Population-Based Prospective Study

**DOI:** 10.64898/2026.01.02.26343366

**Authors:** Ziyang Zheng, Jiaqi Wu, Qilin Li, Xiyuan Qin

## Abstract

**Background:** The relationship between functional tooth units (FTU) and cardiovascular diseases (CVD) risk remains understudied. This study is to investigate the association between masticatory performance, measured by FTU, and CVD incidence and CVH, and mortality among individuals with CVD using data from the National Health and Nutrition Examination Survey (NHANES).

**Methods:** Masticatory function was measured by FTU, CVD was determined by the self-reported questionnaire, CVH was assessed using the American Heart Association’s Life’s Simple 7 (LS7), Life’s Essential 8 (LE8), and Life’s Crucial 9 (LC9). CVD mortality was determined from the National Death Index. Weighted logistic, linear regression and cox proportional hazards models were used to evaluate the associations between FTU and CVD incidence, CVH and mortality.

**Results:** Higher FTU was associated with a reduced risk of CVD. Participants with FTU scores of 9-12 had a 46% reduced risk of CVD compared to those with FTU scores of 0-3. FTU was also positively correlated with better CVH scores in LS7, LE8 and LC9. In the mortality analysis, those with FTU of 9-12 was associated with a 42%, 39% and 71%% reduction in all-cause, CVD- and cancer related mortality. The other masticatory function parameters and sensitivity analyses confirmed these relationships.

**Conclusion:** Preservation of masticatory function, measured by FTU, is associated with decreased CVD prevalence, improved CVH and reduced mortality in individuals with CVD. These findings suggest that the maintenance of optimal masticatory function may play a protective role in reducing CVD incidence and mortality.

**Clinical relevance:** *What is New?:* This study provides novel insights into the association between masticatory function, assessed by functional tooth units (FTU), and cardiovascular disease (CVD) outcomes in individuals with CVD. It demonstrates that higher FTU scores are linked to a significantly lower risk of CVD, improved cardiovascular health (CVH) scores, and reduced mortality, including CVD- and cancer-related deaths. Importantly, this is one of the first studies to show the potential role of masticatory function as a modifiable factor in cardiovascular health and mortality reduction in a large, nationally representative cohort.

*What Are the Clinical Implications?:* The findings highlight the importance of maintaining masticatory function as part of a comprehensive approach to CVD prevention and management. Clinicians should consider the role of oral health, specifically masticatory function, in improving cardiovascular health outcomes and reducing mortality risk in CVD patients. These results suggest that enhancing masticatory function may be a simple, yet effective intervention for reducing CVD incidence and mortality, emphasizing the need for oral health preservation in clinical practice, particularly among individuals with cardiovascular disease.

## Introduction

Cardiovascular diseases (CVD), including coronary heart disease, heart failure, heart attack, angina, and stroke, are the leading cause of global mortality and a major contributor to disabilities. The number of cases has more than doubled, rising from 271 million in 1990 to 523 million in 2019.^1^ From 2025 to 2050, the projected increase in cardiovascular disease prevalence is 90%, with crude mortality expected to rise by 73.4%. Ischemic heart disease will remain the leading cause of cardiovascular deaths.^2^ Similarly, the annual health care cost are projected to almost quadruple and the productivity loss are to increase by 54%.^3^

Emerging evidence have disclosed the bi-directional relationships between chronic periodontitis and atherosclerotic vascular disease, and those with CVD have higher incidence of caries and tooth loss.^4^ Among these oral diseases, the associations between periodontitis and systemic diseases have been widely established, specifically pertaining to cardiovascular diseases and associated diagnoses.^5,6^ As the first step to well-being, the maintenance of overall oral health presents as a modifiable factor for the prevention and treatment of CVD.^7^

Masticatory capacity (chewing ability) is not only a key component of oral health, but is also closely associated with overall health status. Both objective masticatory function and patients’ subjective perception of their chewing performance have been linked to oral-health-related quality of life and general well-being. ^8,9^ Patients with lowest numbers of chewable foods were reported to be associated with higher risks of CVD mortality in active elderly individuals, ^10^ which may be influenced by nutritional pathway with higher level of IL-6 and lower of NTproBNP.^11^

Lower masticatory performance has been reported to be a risk factor for the development of metabolites syndrome and its components including high blood pressure, high triglycerides, and high fasting plasma glucose in Japanese.^12^ Similar, the lower maximum bite force as an objective and quantitative index of masticatory function have been reported to a risk factor of CVD development.^13^

Moreover, preserved masticatory function as measured by the functional tooth units (FTU) has been reported to be with cardiovascular health (CVH) as measured by the American Heart Association Life’s Simple 7 (LS7) in Paris.^14^ The new metric, Life’s Crucial 9 (LC9), built upon LS7 by incorporating sleep and psychological health ^15^, provided a comprehensive evaluation of CVH and was related to a reduced risk of CVD mortality.^16^ The masticatory function have be correlated with cognition, depression and all-cause and CVD mortality.^17–19^

However, the associations between masticatory performance, as measured by FTU, and CVH, CVD incidence, and its mortality in CVD individuals have been less extensively investigated among different population. Therefore, this study is to investigate the associations between masticatory function measured by FTU and the CVH measured by LC9 and its impact on the mortality in the CVD patients from a large sample.

## Methods

### Data origin

This study is utilized 10 cycles based on the National Health and Nutrition Examination Survey (NHANES) from 1999 to 2018. NHANES is conducted by the National Center for Health Statistics (NCHS) and aims to provide a comprehensive assessment of the nutritional and health status of the U.S. population. The Ethics Review Committee of the NCHS revised and approved the protocol of NHANES 1999 to 2018 and written informed consent was obtained from all participants.

The data screening workflow is illustrated in Figure 1, which includes a total of 37,464 participants participated between 1999 to 2018. Among this population, 27849 and 26666 participants had records related to FTU information about lymphocyte and hs-CRP, and 17544 underwent a PHQ questionnaire with validate response. After applying strict covariate inclusion criteria, the final sample size was reduced to 12,868 participants. During this process, missing data were presented in Table 1, with all missing proportions being less than 5%. Therefore, data imputation was not performed.

**Figure 1:**
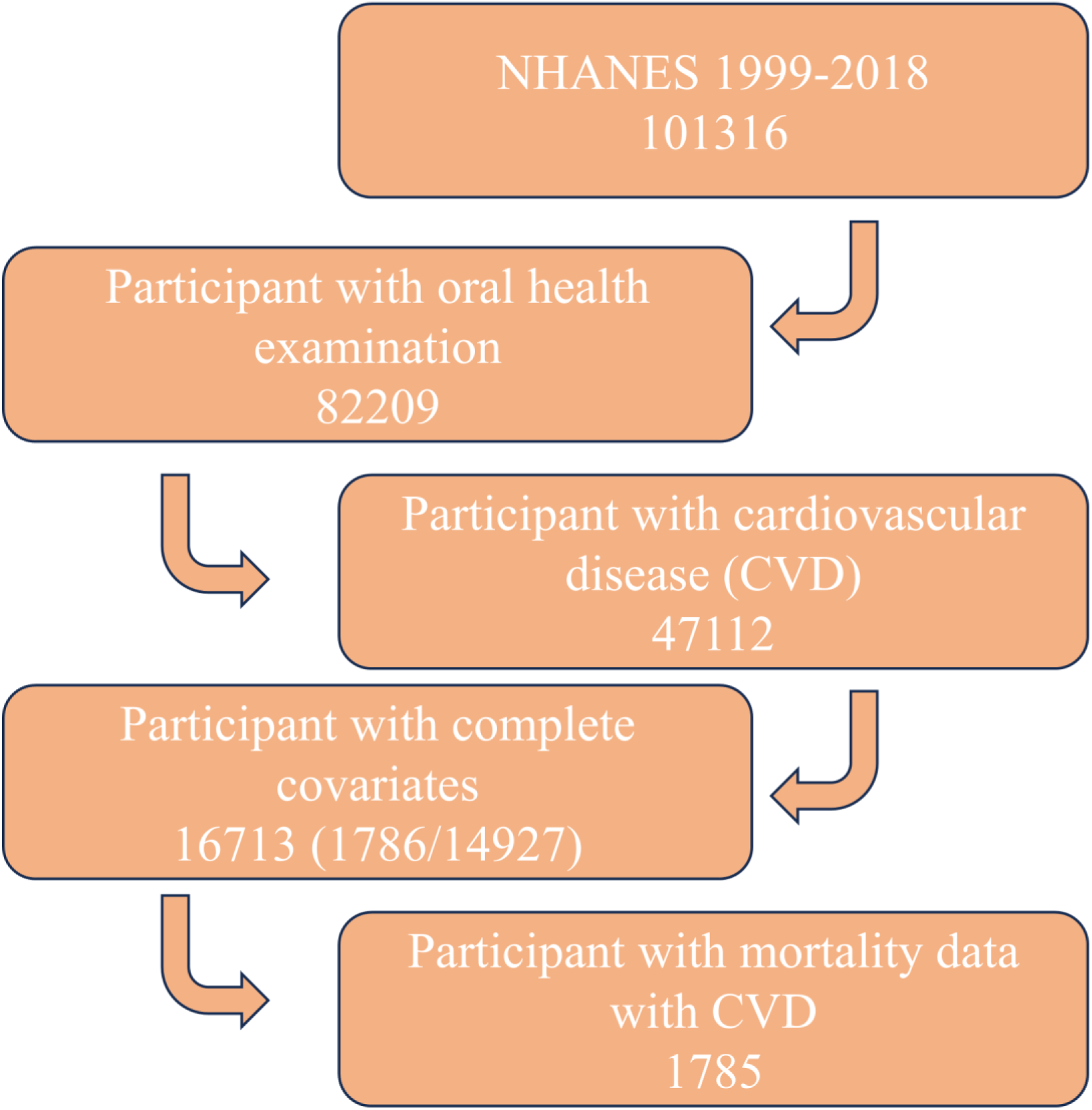
The flowchart of this study.

**Figure 2:**
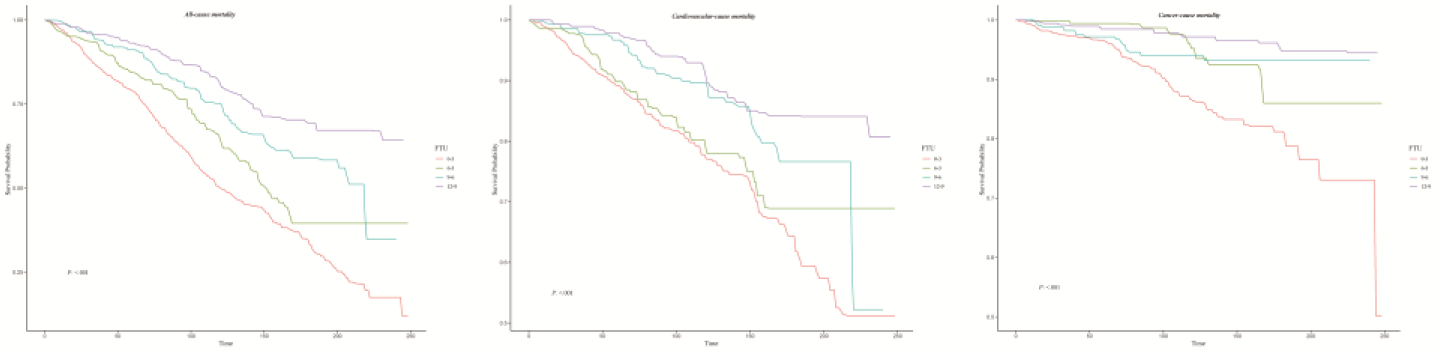
Kaplan-Meier survival analysis for FTU categories and cause-specific mortality. Abbreviation: FTU, Functional tooth units; CVD, cardiovascular diseases.

**Table 1:** Baseline characteristics by the presence of CVD.

| Variable | Total | Without CVD | With CVD | *P*-value |
| --- | --- | --- | --- | --- |
| Age | 46.99(0.24) | 45.39(0.24) | 64.08(0.43) | <.001 |
| Left teeth | 23.19(0.13) | 23.83(0.11) | 16.37(0.39) | <.001 |
| FTU | 8.35(0.08) | 8.71(0.07) | 4.56(0.18) | <.001 |
| Triglycerides (mg/L) | 120.80(0.87) | 119.08(0.84) | 139.09(2.39) | <.001 |
| Total cholesterol (mg/L) | 194.71(0.48) | 195.80(0.51) | 183.08(1.44) | <.001 |
| HDL (mg/L) | 54.06(0.21) | 54.32(0.21) | 51.26(0.59) | <.001 |
| LDL (mg/L) | 116.11(0.40) | 117.16(0.42) | 104.96(1.17) | <.001 |
| Gender |  |  |  | <.001 |
| Female | 8490(51.23) | 7752(51.94) | 738(43.63) |  |
| Male | 8223(48.77) | 7175(48.06) | 1048(56.37) |  |
| Race |  |  |  | <.001 |
| Mexican American | 2862(7.56) | 2674(7.92) | 188(3.67) |  |
| Non-Hispanic Black | 3179(10.44) | 2820(10.38) | 359(11.10) |  |
| Non-Hispanic White | 7964(70.73) | 6918(70.16) | 1046(76.80) |  |
| Other Hispanic | 1293(5.03) | 1183(5.20) | 110(3.24) |  |
| Other Race | 1415(6.24) | 1332(6.34) | 83(5.19) |  |
| Education level |  |  |  | <.001 |
| Below high school | 4121(16.02) | 3523(15.13) | 598(25.44) |  |
| High school | 3843(23.94) | 3405(23.61) | 438(27.43) |  |
| Above high school | 8749(60.05) | 7999(61.26) | 750(47.13) |  |
| Poverty |  |  |  | <.001 |
| Low income | 4753(19.70) | 4152(19.12) | 601(25.78) |  |
| Moderate income | 6482(36.44) | 5724(35.86) | 758(42.53) |  |
| High income | 5478(43.87) | 5051(45.01) | 427(31.69) |  |
| Marital status |  |  |  | <.001 |
| Married/Living with Partner | 10419(65.51) | 9362(65.56) | 1057(64.96) |  |
| Widowed/Divorced/Separated | 3601(17.78) | 2970(16.61) | 631(30.22) |  |
| Never married | 2693(16.71) | 2595(17.82) | 98(4.83) |  |
| Body mass index |  |  |  | <.001 |
| Normal | 4916(31.32) | 4531(32.41) | 385(22.53) |  |
| Overweight | 5654(33.14) | 5059(33.40) | 595(33.58) |  |
| Obese | 5971(34.72) | 5219(34.19) | 752(43.89) |  |
| Smoke |  |  |  | <.001 |
| Never | 9026(53.44) | 8338(55.01) | 688(36.63) |  |
| Former | 4358(25.82) | 3631(24.45) | 727(40.38) |  |
| Now | 3329(20.74) | 2958(20.53) | 371(22.99) |  |
| Alcohol consumption |  |  |  | <.001 |
| Never | 2304(10.87) | 2060(10.83) | 244(11.39) |  |
| Former | 2943(14.45) | 2374(13.02) | 569(29.71) |  |
| Mild | 5814(37.32) | 5181(37.16) | 633(38.96) |  |
| Moderate | 2448(17.00) | 2290(17.74) | 158(9.11) |  |
| Heavy | 3204(20.35) | 3022(21.25) | 182(10.83) |  |
| Diabetes |  |  |  | <.001 |
| No | 13884(88.18) | 12690(91.09) | 1194(70.87) |  |
| Yes | 2362(10.53) | 1772(8.91) | 590(29.13) |  |
| Hypertension |  |  |  | <.001 |
| No | 9705(63.05) | 9280(66.52) | 425(26.31) |  |
| Yes | 7005(36.92) | 5644(33.48) | 1361(73.69) |  |
| Functional dentition |  |  |  | <.001 |
| No | 4022(17.32) | 3067(14.49) | 955(47.40) |  |
| Yes | 12691(82.68) | 11860(85.51) | 831(52.60) |  |
| Masticatory dysfunction |  |  |  | <.001 |
| No | 11588(77.42) | 10916(80.55) | 672(44.15) |  |
| Yes | 5125(22.58) | 4011(19.45) | 1114(55.85) |  |

Abbreviation: FTU, functional tooth unit; HDL, high-density lipoprotein cholesterol; LDL, low-density lipoprotein cholesterol.
Missing data: Body mass index (n=172, 1.03%), Diabetes (n=467, 2.79%), Hypertension (n=3, 0.02%).

### The definition of masticatory performance

The masticatory performance is measured by three aspects including FTU, functional dentition and the number of teeth. Defined as pairs of opposing teeth (including both natural and artificial teeth) in the posterior region (excluding third molars), FTUs were counted as 1 for premolars and 2 for molars, with a total score from 0 to 12, and the tooth position and mobility were not taken into account.^20^ FTU were further categorized into 0-3, 4-6, 7-9, and 9-12.^17^ Impaired mastication was defined as the FTU less than 5.^21^ Functional dentition is defined as having ≥20 natural permanent or prosthetic teeth, while unfunctional dentition is defined as having fewer than 19 teeth.^22^

### The definition of CVH including LS7, LE8 and LC9

CVH is measured by 3 indexes developed by American heart association through modifiable health behaviors and clinical factors including LS7, LE8 and LC9. LS7 is measured by seven components: smoking status, body mass index (BMI), physical activity, total cholesterol (TC), blood pressure, fasting blood glucose (FBG), and diet quality.^23^ LE8 adds the sleep health measured by average sleep hours per night based on LS7.^24^ Then, LC9 adds the psychological health of depression severity measured by Patient Health Questionnaire-9 (PHQ-9).^15^ Each component of the nine scales is standardized to a score of 100, and the overall score is then calculated to represent the CVH status.

### The diagnosis of CVD

The CVD status was determined based on self-reported physician diagnoses and standardized medical history questionnaires completed during individual interviews. Participants who answered “Yes” to any of the following conditions—coronary heart disease, congestive heart failure, heart attack, angina, or stroke—were classified as having CVD.

### Ascertainment of mortality

The mortality data were obtained from National Death Index death certificate records through December 31, 2019. The follow-up time was defined as the interval from the interview date to the date of mortality or through December 31, 2019 for alive participants. The underlying cause of mortality was identified based on the International Classification of Diseases, Tenth Revision. All-cause mortality was defined as death from any cause during the follow-up period. CVD mortality included deaths attributed to heart and cerebrovascular diseases (ICD-10: I00–I09, I11, I13, I20–I51, I60–I69). Cancer mortality was defined as deaths with ICD-10 codes C00–C97.

### The inclusion of covariates

The inclusion of covariates was aligned with previous studies^17,25^. This study included demographic factors such as sex (male or female), race (Mexican American, non-Hispanic White, non-Hispanic Black, and other), education level (High school, high school, or >high school), marital status (Married, living with a partner, widowed, divorced, separated, or never married), and poverty income ratio (PIR: low income defined as a PIR less than 1.5, middle income 1.5-3.5, and greater than 3.5 as high income), and lifestyle factors like smoking status (Never, former, or current) and alcohol consumption (Never, former, mild, moderate, or heavy), while clinical factors included diabetes mellitus (DM), determined by FBG ≥7.0 mmol/L, HbA1c ≥6.5%, use of hypoglycemic drugs, or self-reported physician diagnosis; hypertension, determined by antihypertensive medication use, systolic blood pressure ≥140 mmHg or diastolic blood pressure ≥90 mmHg, or prior physician diagnosis. Moreover, physical and laboratory variables included obese status measured by the body mass index (BMI: normal defined as a BMI less than 25, overweight 25 to 30, and greater than 30 as obese), tooth counts, and FTU.

### Statistical analyses

In compliance with NHANES analytical guidelines, this study recalculated the 20-year composite weights by 4/20 of the 4-year medical examination center weights from NHANES 1999-2002, added with the 2/20 of the 2-year medical examination center weights from NHANES 2003-2018. The basic characteristics of the survey included participants were stratified into two groups based on the presence or absence of CVD and mortality status. Continuous variables were expressed as mean± standard deviation (SD), whereas categorical variables as proportions (%) and counts (n). Weighted Student’s t-tests and weighed Chi-square tests were used to compare baseline characteristics.

Multiple logistic and linear regression models were used to calculate the odds ratio (OR), beta coefficient (β) and confidence interval (CI) to assess the impact of masticatory performance related exposures on CVH and CVD, Cox proportional hazards model was used to assess the associations between masticatory performance and mortality in participants with CVD. The proportional risk assumption was examined using Schoenfeld residuals. In the mortality study, a sensitivity analysis was performed by excluding participants who died within the first two years.

Three models were established: Model I (No adjustment), Model II (Adjustment for age, sex, race, poverty, marital status and education level) and Model III (Adjustment for age, sex, race, poverty, marital status, education level, BMI, smoke, alcohol consumption, diabetes, hypertension).

Subgroup analyses were conducted. Restricted cubic spline (RCS) analysis was conducted to determine whether a non-linear relationship existed between FTU and CVD and mortality. All analyses accounted for the survey design and weighting variables using R software (version 4.3.3). A *P*-value less than 0.05 was considered statistically significant.

## Results

### Basic characteristics

The data screening flowchart is presented in Figure 1, with a total of 16,713 in the cross-sectional study and 1,785 in the prospective study. The baseline of included participants by the presence of CVD is shown in Table 1, and in Table S1 by the cause-specific mortality. The baseline of the cross-sectional design by the FTU categories are details in Table S2. In the cross-sectional study, 1,786 were reported CVD with a weighted prevalence of 8.58%, those with and without CVD had a FTU score of 4.56 and 8.71 respectively, and a number of teeth of 16.37 and 23.83 respectively, with a significant difference. Moreover, those with CVD were more likely to have masticatory dysfunction. In this prospective study, 1,785 participants were included, among whom 771 deaths occurred, including 336 from CVD and 139 from cancer. Notably, those who died had FTU scores below 3 and the number of teeth less than 13.

### Associations between masticatory function and CVD

The relationship between masticatory function measured by FTU and CVD is presented in Table 2, both continuous and categorical FTU were associated with decreased risk of CVD. After adjusting all the covariates, each unit in FTU was associated with a 6% decrease of CVD development (OR: 0.94, CI: 0.92,0.97; *P*<.001); compared with FTU of 0-3, those with FTU 9-12 had a 46% decrease risk of CVD (OR: 0.54, CI: 0.42,0.69; *P*<.001). Higher FTU was associated with reduced odds of CVD (*P* for trend <0.05). The correlation between masticatory function parameters and CVD is presented in Table S3, premolar and molar FTU, tooth count, and functional dentition were associated with lower risk of CVD while masticatory dysfunction was with increased risk of CVD in the fully adjusted model. These results unveiled that masticatory function were significantly associated with CVD incidence.

**Table 2:**
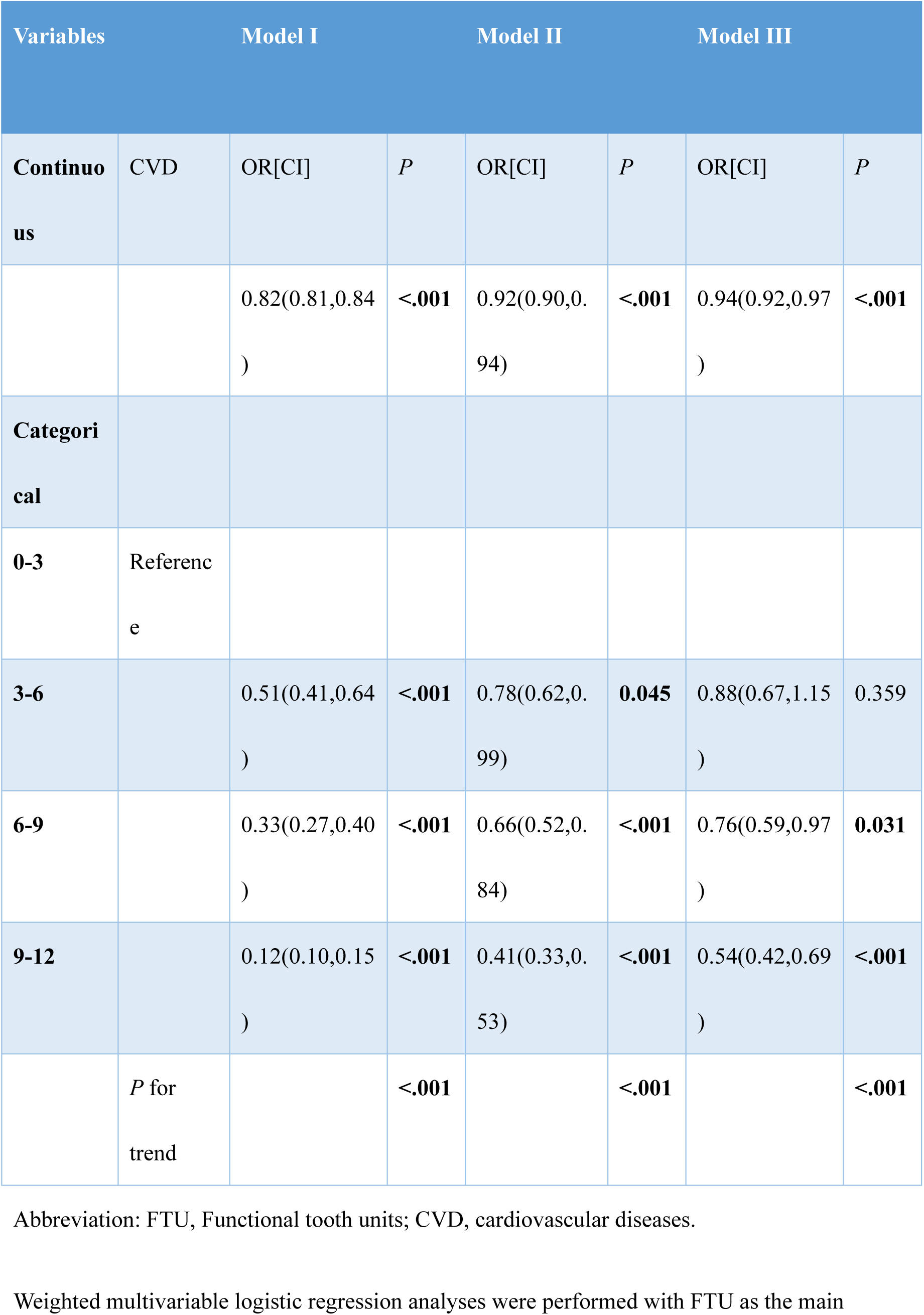
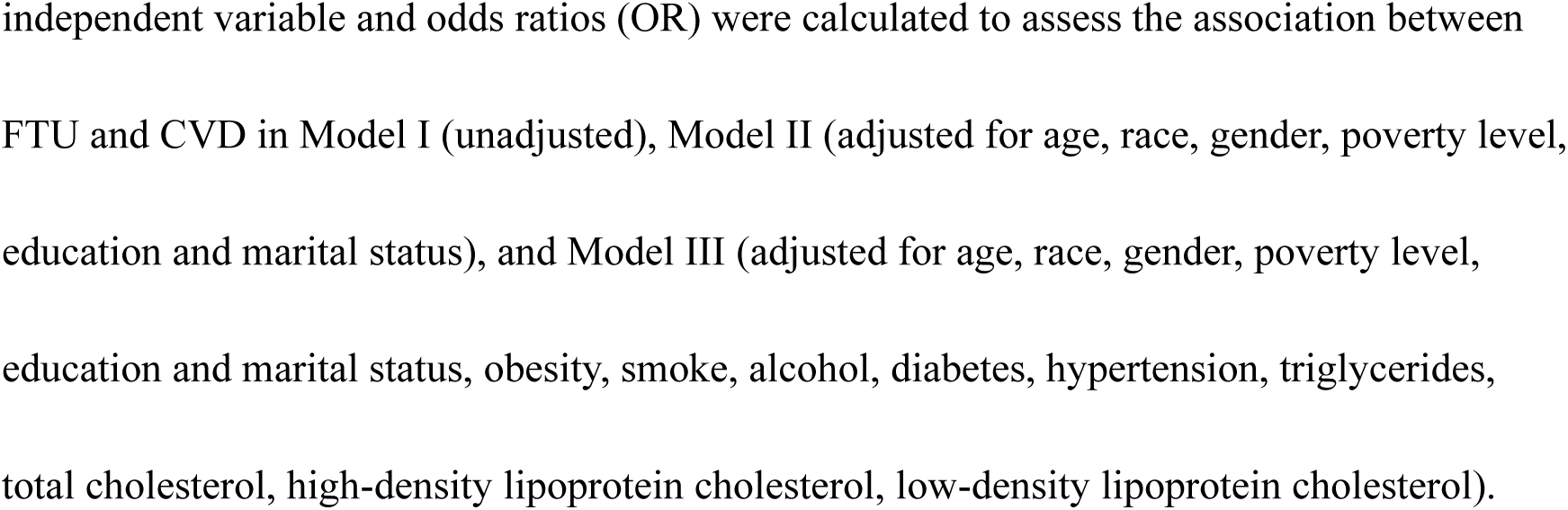
Association between FTU and CVD.

**Table 3:**
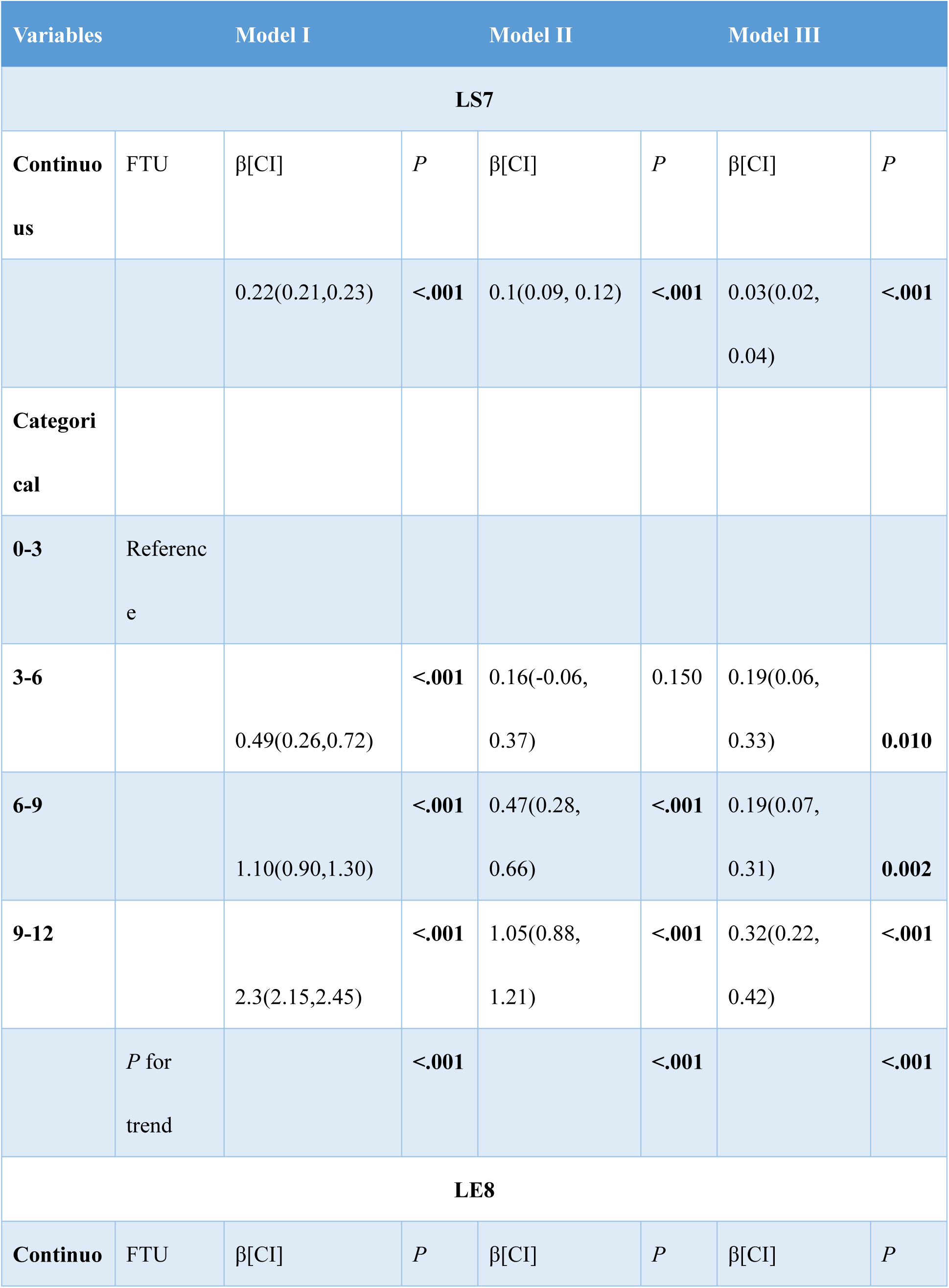
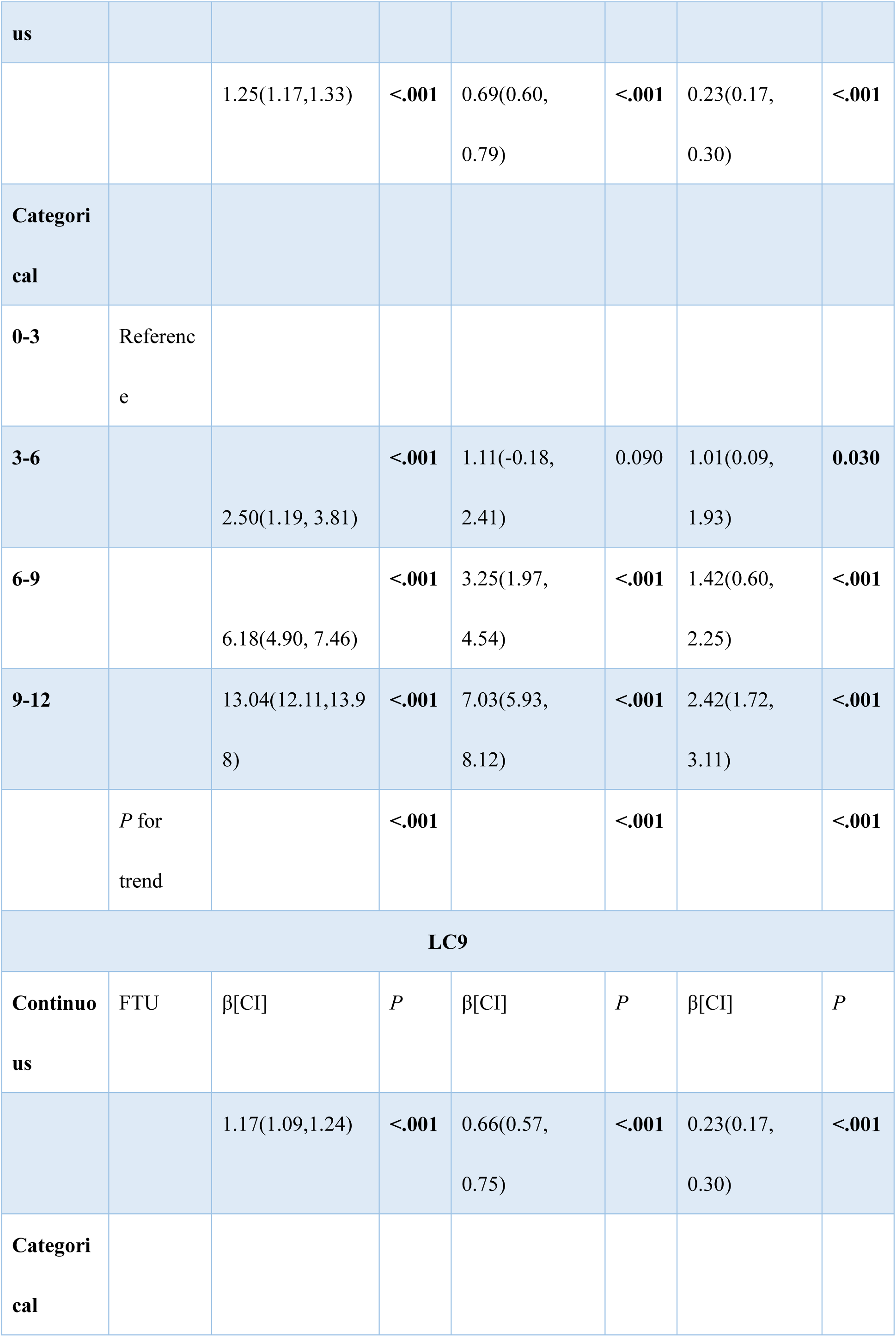
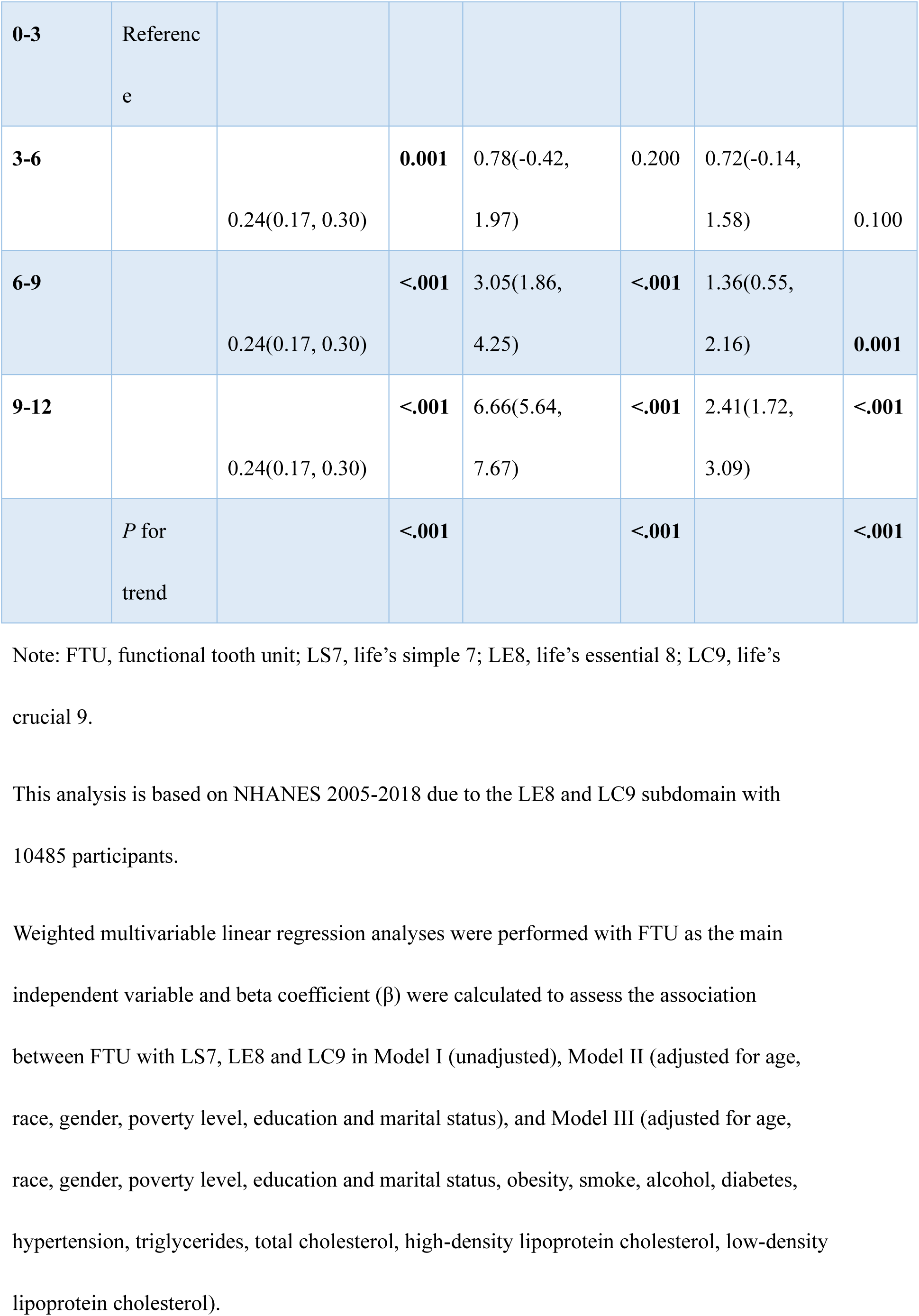
Association between FTU with LS7, LE8 and LC9.

### Associations between masticatory function and CVH

To further delve the relationship between masticatory function and CVH, the relationship between FTU and CVH measured by LS7, LE8 and LC9 is depicted in Table 4. FTUs in continuous and categorical were all associated with increased score of LS7, LE8 and LC9, and higher FTU was associated with elevated CVH score (*P* for trend <0.05), revealing that FTU was associated with cardiovascular function. The associations between masticatory parameters and CVH are presented in Table S4. After adjustment for all covariates, premolar and molar FTU, tooth count, and functional dentition were associated with lower CVH scores, whereas masticatory dysfunction was associated with higher CVH scores. These findings highlighted that masticatory function may contribute to reduced CVD prevalence by enhancing CVH.

**Table 4:**
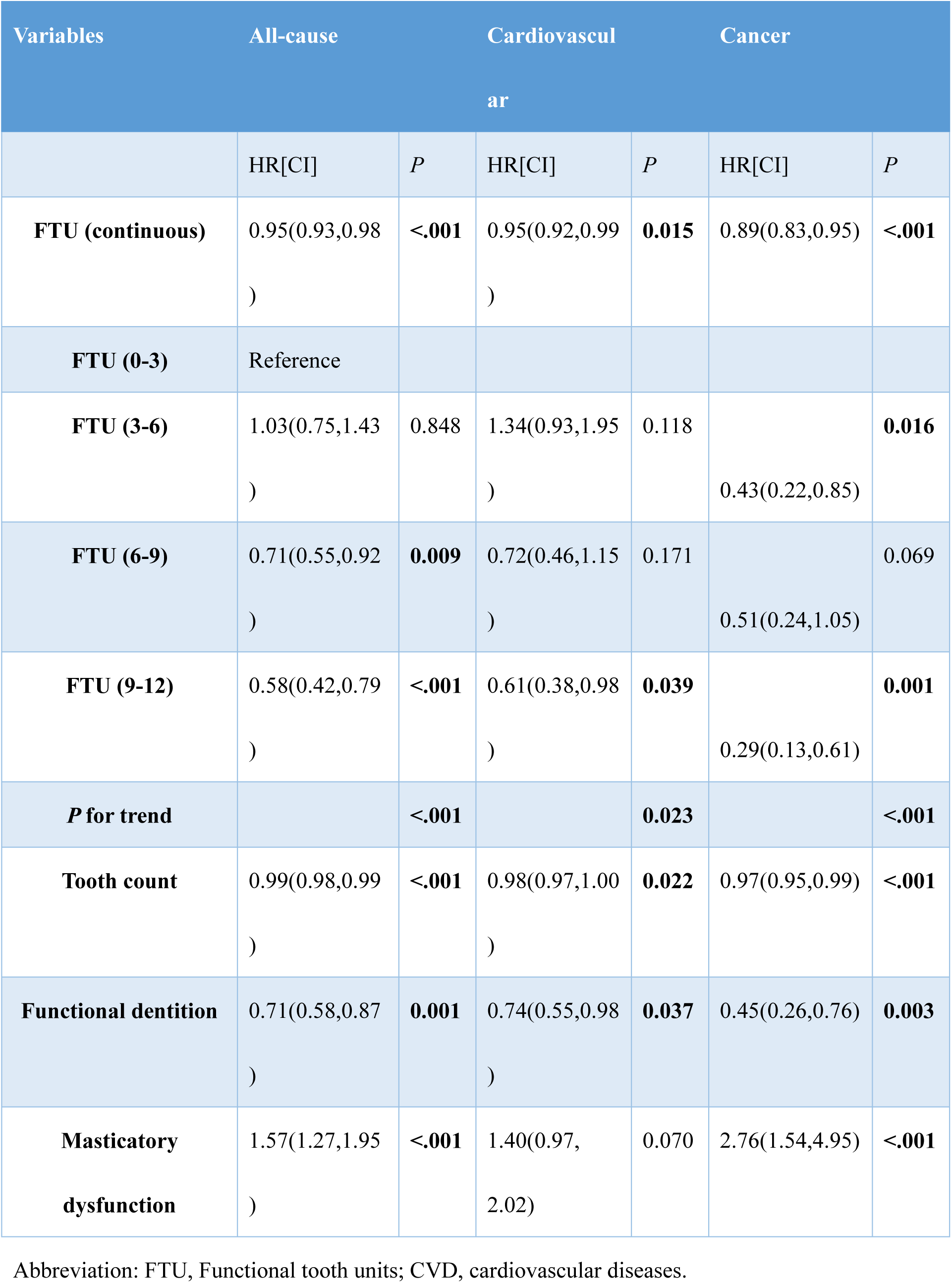
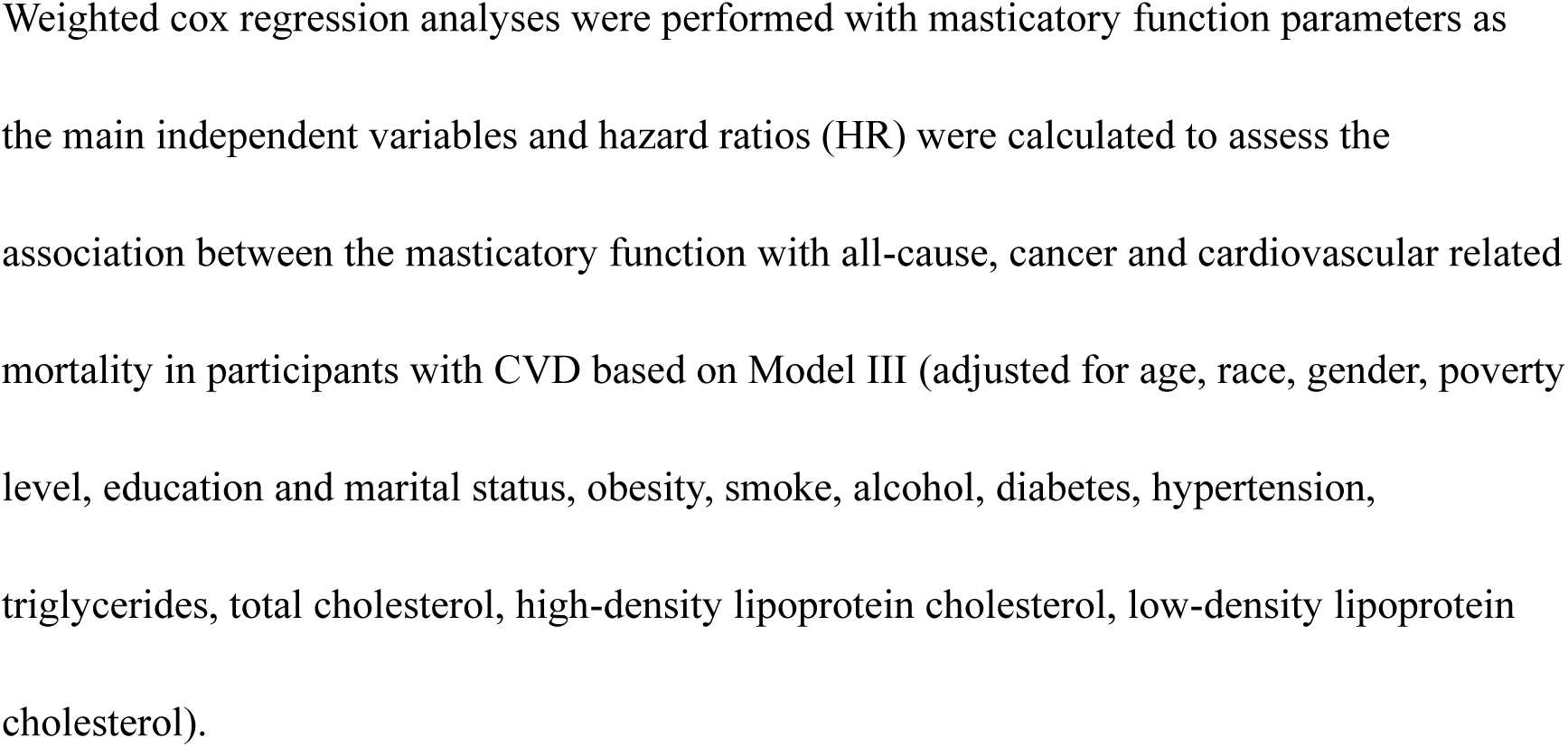
Association of FTU and masticatory indicators with the all and cause-specific mortality in participants with CVD.

### Associations between masticatory function and mortality among those with CVD

The Kaplan-Meier survival analyses by FTU subgroups are displayed in Figure 3, revealing that those with 9-12 score of FTU had the highest survival. The results of masticatory function and cause-mortality in those participants with CVD are presented in Table 5. Each one-unit increase in FTU was associated with a 5% reduction in all-cause mortality (HR: 0.95; 95% CI: 0.93–0.98; *P*<.001), a 5% reduction in CVD-cause mortality (HR: 0.94; 95% CI: 0.92–0.99; *P*=0.015), and an 11% reduction in cancer- cause mortality (HR: 0.89; 95% CI: 0.83–0.95; *P*<.001). Subjects with FTU scores of 9-12 had a 42%, 39%, and 71% reduction in all-cause, CVD-related, and cancer-related mortality, respectively, compared to those with FTU scores of 0-3. Also, higher FTU was associated lower mortality in those with CVD (*P* for trend <0.05). Tooth count and functional dentition were associated with reduced mortality, while masticatory dysfunction was associated with increased mortality, except for CVD-related mortality. Sensitivity analyses, excluding individuals who died within the first 2 years of follow-up, are presented in Table S5. FTU, tooth count, and functional dentition were negatively associated with mortality, while masticatory dysphagia was positively associated with mortality. These findings suggest that masticatory function is linked to lower mortality in individuals with CVD.

**Figure 3:**
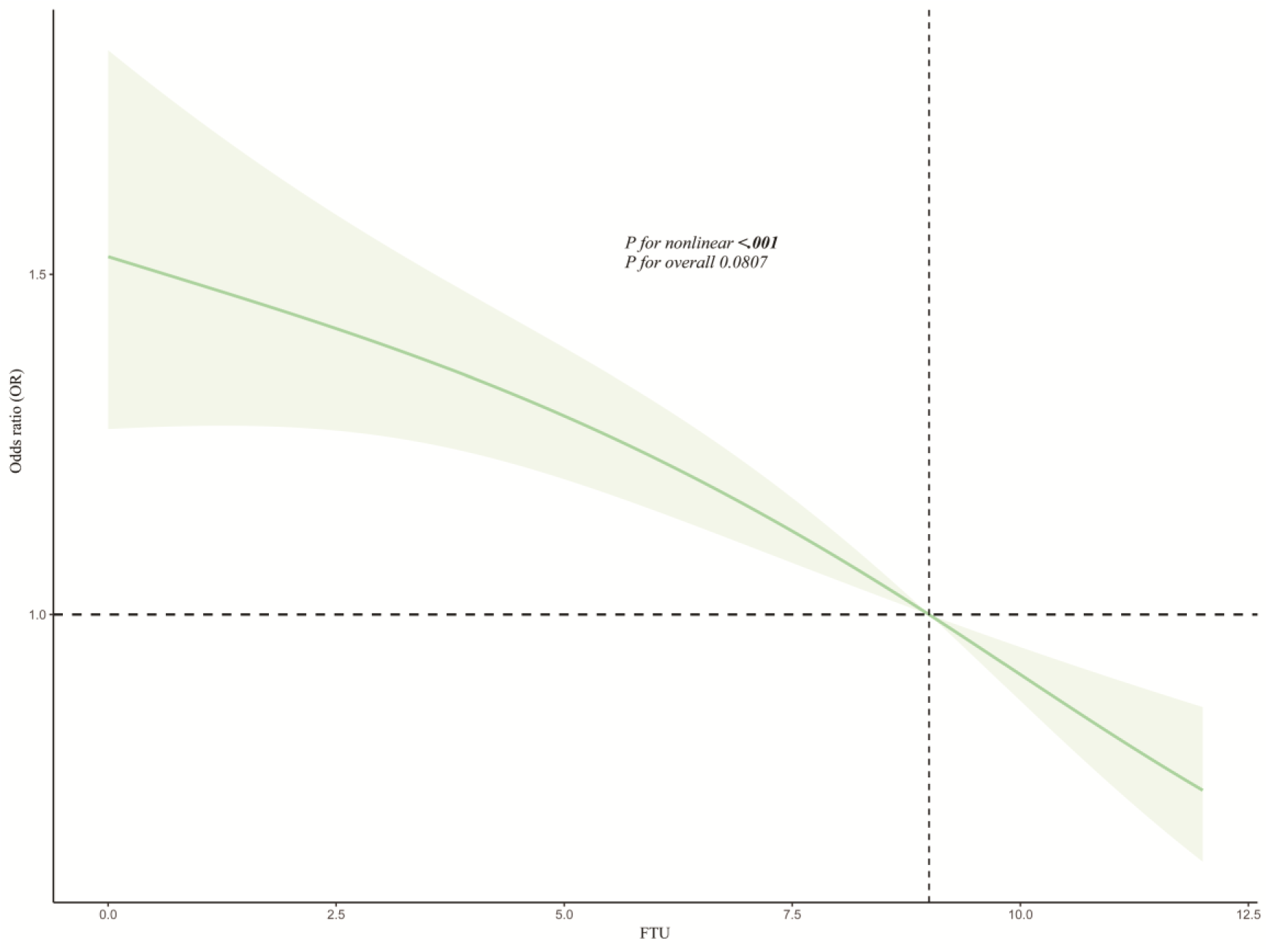
RCS plot of relationship between FTU and CVD. Abbreviation: FTU, Functional tooth units; CVD, cardiovascular diseases.

### RCS and stratified analyses

To explore the linear relationship between FTU and CVD, CVH, and mortality in individuals with CVD, RCS analyses were conducted. In the relationship between FTU and CVD (Figure 3), a linear curve was observed (*P* for non-linearity > 0.05). The RCS curve for FTU and CVH, as measured by LS7, LE8, and LC9 (Figure S1), revealed a non-linear relationship between FTU and LS7 (*P* for non-linearity = 0.0175), while linear relationships were observed for LE8 and LC9 (all P for non-linearity > 0.05). Regarding the relationship between FTU and cause-specific mortality (Figure 4), a non-linear relationship was found for all-cause mortality (*P* for non-linearity = 0.0079), while linear relationships were identified for CVD and cancer-related mortality (all *P* for non-linearity > 0.05). These results indicate that higher FTU is associated with a lower risk of CVD, higher CVH scores, and reduced mortality in individuals with CVD.

**Figure 4:**
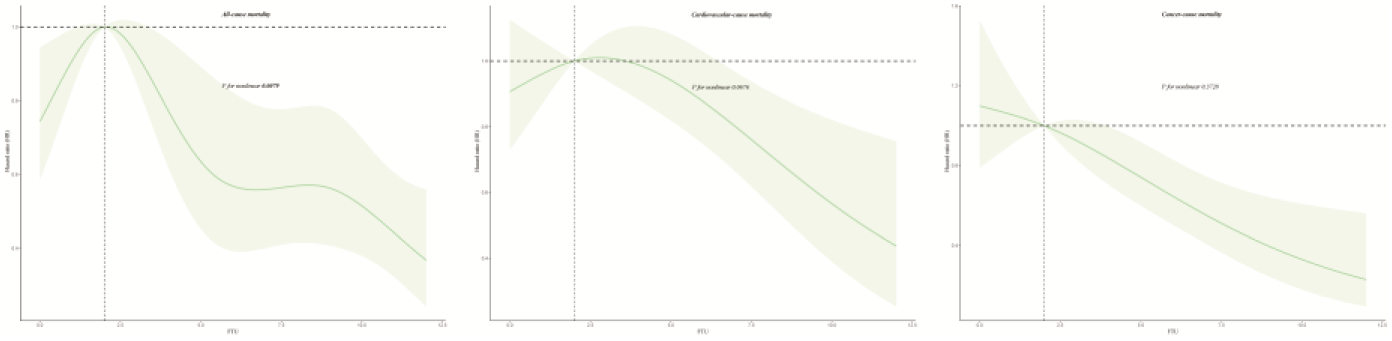
RCS plot of relationship between FTU and cause-specific mortality. Abbreviation: FTU, Functional tooth units; CVD, cardiovascular diseases.

Subgroup analyses were also conducted. Age and poverty were found to interacted with FTU in relation to CVD risk (Figure S2), with stronger ORs observed in younger individuals and those with middle incomes. The subgroup analysis of FTU on mortality (Figure S3) revealed that age and smoking status interacted with FTU in relation to cancer mortality, with stronger HRs in younger individuals and current/former smokers.

## Discussion

To the authors known, this study is the first study to investigate the association between FTU and CVH, CVD incidence and mortality with CVD. Based on this U.S. large representative sample, this study revealed that better masticatory function was associated with higher CVH score measured by LC9, lower prevalence of CVD and decreased incidence of all-cause and CVD related mortality of CVD patients.

Oral health is an importance segment of the general health, the impaired oral status is associated with mastication and subsequent nutritional problems.^26^ Poor oral health is associated not only with negative outcomes in general health, but also with impairments in psychological, social, and cognitive function. The dental health combining plaque and oral health score were significantly with correlated with fatal coronary events.^27^ Moreover, in those patients with CVD excluded, poor oral health was associated with an increased mortality risk.^28^

Also, as an important part of the oral health, the mastication is affected by almost all investigated characteristics, including age, socioeconomic status, physical activities, depressive symptoms, smoking, dental facilities utilization and the needs for dental care.^29^ Additionally, the cognitive impairment, the check-up history of medical visit, the number of chronic diseases could predict the chewing ability.^30^ Several oral diseases including the dry mouth, tooth loss, caries have also been identified to be associated with the impact oral function.^31^

The measurements of mastication could be measured by the objective methods including the colorable chewing gum, maximum bite forces, the oral health behaviors, and the subjective methods including the self-reported masticatory ability, the number of foods kinds be chewed and dietary hardness.^32^ However, the use of standardized measuring devices, self-reported recall bias, and variations in dietary habits may limit the accuracy of masticatory function assessment. The number of teeth, functional tooth pairs, prosthetic dental status, and periodontal condition are all associated with masticatory performance.^33^ Hence, the FTU have been introduced into clinical practice due to their convenience and feasibility.

In the patients with periodontitis, the number of teeth and depended pockets were significantly with future MI and HF in Thailand,^34^ which may be mediated by higher levels of C-reactive protein (CRP), fibrinogen, factor VII, tissue plasminogen activator (t-PA), LDL-C, IL-6 and neutrophils.^35^ Moreover, periodontitis Grade B/C was associated with higher CVD risk than no periodontitis/Grade A in Norway.^36^ The PAROKRANK study also revealed that CVD events were more common among participants with periodontitis.^37^ Similarly, the participants with community periodontal index 4 have higher systemic coronary risk estimation (SCORE) values.^38^ Also, it was reported that the severity of coronary artery stenosis in Chinese old patients was positively with the severity of periodontitis.^39^ Similarly, based on a large Korean cohort, there was a dose-dependent relationship between tooth loss and MI, HF, ischemic stroke and all-cause death, and could serve as a good predictor of cardiovascular outcomes.^40^ Also, tooth loss levels were associated with CVD in Serbia.^41^ It is noted that there is a causal effect of tooth loss on CVD among US adults.^42^ Moreover, the apical periodontitis is an independent and important marker of cardiovascular vulnerability, ^43^ which is mediated by the impaired flow-mediated dilatation and carotid intima-media thickness between endodontic infection.^44^ In addition, the higher decayed, missing and filled teeth (DMFT index) scores and missing teeth were positively correlated with CVD incidence.^45^ Self reported gingivitis, dental caries and tooth loss were also associated with angina pectoris.^46^ Furthermore, dental caries was also independently with the ischemic stroke and death.^47^

Patients with CVD are showing the paucity of oral health education.^48^ Meantime, previous study revealed that there was an positive association between FTU and ideal CVH in French,^14^ which was aligned with this research. Conversely, in the associations between CVH and oral health, LE8 and LC9 were both positively associated with periodontitis.^49,50^

Several mechanisms maybe under the associations between mastication and CVH and CVD incidence. The mechanisms could be the nutrition, systemic inflammation and the hemodynamic changes.

There is extensive evidence demonstrating that dietary interventions are effective in reducing CVD incidence and improving the treatment of CVD.^51^ Impaired masticatory function can hinder the consumption of foods, particularly those with harder textures, and the provision of prosthetic treatment and dietary interventions can lead to changes in food intake.^52^ Moreover, lower masticatory performance was associated with a lower intake of vitamin A, β-carotene and folic acids in the older Japanese.^53^ Impaired mastication may lead to food avoidance and a higher complaints of digestion^54^ In females, the low masticatory ability was associated with low levels of plasma albumin and low BMI, which underlined the preserved masticatory capacity for undernutrition prevention.^55^ Targeting a new dietary option for patients with impaired mastication may provide a new perspective to halt this nutritional pathway.

Moreover, the periodontal bacteria and its products induced bacteremia and sepsis could circulate in the blood vessels, leading to the elevated systemic levels of inflammatory mediators, thrombotic and hemostatic markers.^56^ These mediators included C-reactive protein, interleukins, matrix metalloproteinases.^57^ Intriguingly, studies revealed that patients with coronary artery calcification had been detected the subgingival plaque bacterial complexes in their blood, and in the cardiac tissue from aortic valve surgery.^58,59^ Then, the genetics susceptibility may overlap in both periodontitis and CAD through epigenetics.^60^ To be noted, the masticatory performance has a correlation with the frailty and obesity and glucose metabolism.^61,62^ Also, higher masticatory performance and slow eating could inhibit the incidence of diabetes.^63^ These may further implement the associations between mastication and CVH by the low-grade inflammation.

Furthermore, there are associations between mastication and cerebral hemodynamics, and the cerebral circulation is activated during gum chewing.^64,65^ Also, study revealed the high sympathetic activity group showed a lower masticatory performance, and mastication was correlated with the heart rate variability.^66^ During the chewing tasks, the heart rate and blood pressure were elevated, showing that mastication is associated general circulatory effects.^67^ In addition, the maximum mouth opening during mastication was followed by reductions in blood pressure and heart rate.^68^ The abovementioned studies revealed that mastication may be connected to CVH by general circulation. However, all these postulated mechanisms need further explained and validated.

It is essential to acknowledge that this study does have several limitations. First, although this study reveals a potential protective role of preserved masticatory performance in maintaining ideal CVH, no causal relationship can be established due to its cross-sectional design. Whether the rehabilitation of impaired mastication contributes to improvements in CVH requires confirmation through prospective studies. Second, the diagnosis of CVD and the assessment of suboptimal CVH components in this study rely largely on self-reported questionnaires, which may introduce recall bias and potentially weaken the observed associations. Lastly, although this study included as many confounding variables as possible based on the NHANES dataset to maintain a sufficiently large sample size, residual confounding from unmeasured covariates cannot be ruled out.

## Conclusion

A positive association has been established between masticatory function and CVH, while a protective association has been observed with CVD incidence and lower mortality among patients with CVD. Maintaining masticatory performance may contribute to a lower incidence of CVD and reduced all-cause and CVD-related mortality in individuals with CVD by the preservation of CVH.

## Abbreviations

NHANES: National Health and Nutrition Examination Survey
BMI: Body mass index
CHD: Coronary heart disease
CHF: Chronic heart failure
CKD: Chronic kidney disease
DM: Diabetes mellitus
HA: Heart attack
CVH: Cardiovascular health
CVD: Cardiovascular diseases

## Nonstandard Abbreviations and Acronyms

CVD: cardiovascular diseases
CVH: cardiovascular health
FTU: functional tooth units
LC9: Life’s Crucial 9
LE8: Life’s Essential 8
LS7: Life’s Simple 7
NHANES: National Health and Nutrition Examination Survey)

## Acknowledgements

We appreciate the NHANES open policy and the provided data as well as all participants and staffs in this study.

## Author contributions

Ziyang Zheng: Conceptualization, Methodology, Writing–review & editing, Data curation, Formal analysis. Jiaqi Wu: Conceptualization, Investigation, Software, Writing–original draft. All authors read and approved the final manuscript.

## Funding

This study did not receive any funding or financial support. All work for this article was fully self-funded by the authors, and no acknowledgments are necessary.

## Data availability statement

The findings of this study are supported by data from the NHANES (National Health and Nutrition Examination Survey). The dataset is publicly available and can be accessed directly at the following link: https://wwwn.cdc.gov/nchs/nhanes/.

## Ethics statement

The studies involving human participants were approved by the Ethics Review Board of the National Center for Health Statistics. They were conducted in accordance with local regulations and institutional guidelines.

## Conflict of interest statement

The authors have no conflicts of interest to disclose.

## Notes

### Competing Interest Statement

The authors have declared no competing interest.

### Clinical Trial

NA

### Author Declarations

The studies involving human participants were approved by the Ethics Review Board of the National Center for Health Statistics. They were conducted in accordance with local regulations and institutional guidelines. All participants provided their written informed consent.

## Reference

1. Roth GA, Mensah GA, Johnson CO, Addolorato G, Ammirati E, Baddour LM, Barengo NC, Beaton AZ, Benjamin EJ, Benziger CP, Bonny A, Brauer M, Brodmann M, Cahill TJ, Carapetis J, Catapano AL, Chugh SS, Cooper LT, Coresh J, Criqui M, DeCleene N, Eagle KA, Emmons-Bell S, Feigin VL, Fernández-Solà J, Fowkes G, Gakidou E, Grundy SM, He FJ, Howard G, Hu F, Inker L, Karthikeyan G, Kassebaum N, Koroshetz W, Lavie C, Lloyd-Jones D, Lu HS, Mirijello A, Temesgen AM, Mokdad A, Moran AE, Muntner P, Narula J, Neal B, Ntsekhe M, Moraes de Oliveira G, Otto C, Owolabi M, Pratt M, Rajagopalan S, Reitsma M, Ribeiro ALP, Rigotti N, Rodgers A, Sable C, Shakil S, Sliwa-Hahnle K, Stark B, Sundström J, Timpel P, Tleyjeh IM, Valgimigli M, Vos T, Whelton PK, Yacoub M, Zuhlke L, Murray C, Fuster V, GBD-NHLBI-JACC Global Burden of Cardiovascular Diseases Writing Group. Global Burden of Cardiovascular Diseases and Risk Factors, 1990-2019: Update From the GBD 2019 Study. J Am Coll Cardiol. 2020;76:2982–3021.

2. Chong B, Jayabaskaran J, Jauhari SM, Chan SP, Goh R, Kueh MTW, Li H, Chin YH, Kong G, Anand VV, Wang J-W, Muthiah M, Jain V, Mehta A, Lim SL, Foo R, Figtree GA, Nicholls SJ, Mamas MA, Januzzi JL, Chew NWS, Richards AM, Chan MY. Global burden of cardiovascular diseases: projections from 2025 to 2050. Eur J Prev Cardiol. 2024;:zwae281.

3. Kazi DS, Elkind MSV, Deutsch A, Dowd WN, Heidenreich P, Khavjou O, Mark D, Mussolino ME, Ovbiagele B, Patel SS, Poudel R, Weittenhiller B, Powell-Wiley TM, Joynt Maddox KE, American Heart Association. Forecasting the economic burden of cardiovascular disease and stroke in the United States through 2050: a presidential advisory from the american heart association. Circulation. 2024;150:e89–e101.

4. Dietrich T, Webb I, Stenhouse L, Pattni A, Ready D, Wanyonyi KL, White S, Gallagher JE. Evidence summary: the relationship between oral and cardiovascular disease. Br Dent J. 2017;222:381–385.

5. Hopkins S, Gajagowni S, Qadeer Y, Wang Z, Virani SS, Meurman JH, Krittanawong C. Oral health and cardiovascular disease. Am J Med. 2024;137:304–307.

6. Seymour GJ, Ford PJ, Cullinan MP, Leishman S, Yamazaki K. Relationship between periodontal infections and systemic disease. Clin Microbiol Infect: Off Publ Eur Soc Clin Microbiol Infect Dis. 2007;13 Suppl 4:3–10.

7. Altamura S, Del Pinto R, Pietropaoli D, Ferri C. Oral health as a modifiable risk factor for cardiovascular diseases. Trends Cardiovasc Med. 2024;34:267–275.

8. Brennan DS, Spencer AJ, Roberts-Thomson KF. Tooth loss, chewing ability and quality of life. Qual Life Res: Int J Qual Life Asp Treat Care Rehabil. 2008;17:227–235.

9. Inukai M, John MT, Igarashi Y, Baba K. Association between perceived chewing ability and oral health-related quality of life in partially dentate patients. Health Qual Life Outcomes. 2010;8:118.

10. Ansai T, Takata Y, Soh I, Yoshida A, Hamasaki T, Awano S, Sonoki K, Akifusa S, Fukuhara M, Sogame A, Shimada N, Takehara T. Association of chewing ability with cardiovascular disease mortality in the 80-year-old japanese population. Eur J Cardiovasc Prev Rehabil: Off J Eur Soc Cardiol Work Groups Epidemiol Prev Card Rehabil Exerc Physiol. 2008;15:104–106.

11. Chatzopoulou E, Rangé H, Deraz O, Boutouyrie P, Perier M-C, Guibout C, Thomas F, Andrieu M, Bailly K, Vedie B, Danchin N, Jouven X, Bouchard P, Empana J-P. Poor Masticatory Capacity and Blood Biomarkers of Elevated Cardiovascular Disease Risk in the Community: The Paris Prospective Study III. *Arterioscler, Thromb*, Vasc Biol. 2021;41:2225–2232.

12. Fushida S, Kosaka T, Nakai M, Kida M, Nokubi T, Kokubo Y, Watanabe M, Miyamoto Y, Ono T, Ikebe K. Lower masticatory performance is a risk for the development of the metabolic syndrome: the suita study. Front Cardiovasc Med. 2021;8:752667.

13. Hashimoto S, Kosaka T, Nakai M, Kida M, Fushida S, Kokubo Y, Watanabe M, Higashiyama A, Ikebe K, Ono T, Miyamoto Y. A lower maximum bite force is a risk factor for developing cardiovascular disease: the suita study. Sci Rep. 2021;11:7671.

14. Rangé H, Perier M-C, Boillot A, Offredo L, Lisan Q, Guibout C, Thomas F, Danchin N, Boutouyrie P, Jouven X, Bouchard P, Empana J-P. Chewing capacity and ideal cardiovascular health in adulthood: a cross-sectional analysis of a population-based cohort study. Clin Nutr (edinb Scotl). 2020;39:1440–1446.

15. Gaffey AE, Rollman BL, Burg MM. Strengthening the pillars of cardiovascular health: psychological health is a crucial component. Circulation. 2024;149:641–643.

16. Ge J, Peng W, Lu J. Predictive Value of Life’s Crucial 9 for Cardiovascular and All-Cause Mortality: A Prospective Cohort Study From the NHANES 2007 to 2018. J Am Heart Assoc. 2024;13:e036669.

17. Zheng Z, Xu M, Wang L, Deng Y, Liu Q, Yu K. Masticatory function and cognition in older adults: a population-based study. J Prosthet Dent. 2025;:S0022391325000435.

18. Wei X, Zhang X, Zhang X, Chen R, Tonetti MS, Shi J, Lai H. Association between masticatory function and depression in older adults: Results from NHANES 2009 to 2018. J Clin Periodontol. 2024;51:1458–1465.

19. Du M, Deng K, Yin J, Wu C, Hu S, Guo L, Luo Z, Tonetti M, Tjakkes G-HE, Visser A, Ge S, Li A. Association between chewing capacity and mortality risk: the role of diet and ageing. J Clin Periodontol. 2025;52:695–706.

20. Wu X, Shen J, Zhang X, Liu B, Liu M, Shi J, Qian S, Zong G, Lai H, Yuan C, Tonetti MS. The potential causal path between periodontitis stage diagnosis and vegetable consumption is mediated by loss of posterior functional tooth units and masticatory function. J Clin Periodontol. 2024;51:691–701.

21. Du M, Deng K, Yin J, Wu C, Hu S, Guo L, Luo Z, Tonetti M, Tjakkes GE, Visser A, Ge S, Li A. Association Between Chewing Capacity and Mortality Risk: The Role of Diet and Ageing. J Clinic Periodontology. 2025;:jcpe.14122.

22. Sheiham A, Steele J. Does the condition of the mouth and teeth affect the ability to eat certain foods, nutrient and dietary intake and nutritional status amongst older people? Public health nutrition. 2001;4:797–803.

23. Lloyd-Jones DM, Hong Y, Labarthe D, Mozaffarian D, Appel LJ, Van Horn L, Greenlund K, Daniels S, Nichol G, Tomaselli GF, Arnett DK, Fonarow GC, Ho PM, Lauer MS, Masoudi FA, Robertson RM, Roger V, Schwamm LH, Sorlie P, Yancy CW, Rosamond WD. Defining and setting national goals for cardiovascular health promotion and disease reduction: the american heart association’s strategic impact goal through 2020 and beyond. Circulation. 2010;121:586–613.

24. Lloyd-Jones DM, Allen NB, Anderson CAM, Black T, Brewer LC, Foraker RE, Grandner MA, Lavretsky H, Perak AM, Sharma G, Rosamond W, on behalf of the American Heart Association. Life’s essential 8: updating and enhancing the american heart association’s construct of cardiovascular health: a presidential advisory from the american heart association. Circulation. 2022;146. doi:10.1161/cir.0000000000001078.

25. Zheng Z, Xu M, Xiao K, Yu K. Association between oral microbiome and depression: A population-based study. Journal of Affective Disorders. 2025;379:441–447.

26. Gil-Montoya JA, de Mello ALF, Barrios R, Gonzalez-Moles MA, Bravo M. Oral health in the elderly patient and its impact on general well-being: a nonsystematic review. Clin Interv Aging. 2015;10:461–467.

27. Jansson L, Lavstedt S, Frithiof L, Theobald H. Relationship between oral health and mortality in cardiovascular diseases. J Clin Periodontol. 2001;28:762–768.

28. Jansson L, Lavstedt S, Frithiof L. Relationship between oral health and mortality rate. J Clin Periodontol. 2002;29:1029–1034.

29. Krause L, Seeling S, Schienkiewitz A, Fuchs J, Petrakakis P. Chewing ability and associated factors in older adults in Germany. Results from GEDA 2019/2020-EHIS. BMC Oral Health. 2023;23:988.

30. Park K, Hong G-RS. Predictors of chewing ability among community-residing older adults in Korea. Geriatr Gerontol Int. 2017;17:78–84.

31. Khalifa N, Allen PF, Abu-bakr NH, Abdel-Rahman ME. Chewing ability and associated factors in a sudanese population. J Oral Sci. 2013;55:349–357.

32. Maekawa K, Motohashi Y, Igarashi K, Mino T, Kawai Y, Kang Y, Hirai T, Kuboki T. Associations between measured masticatory function and cognitive status: A systematic review. Gerodontology. 2024;41:452–463.

33. Fan Y, Shu X, Leung KCM, Lo ECM. Associations of general health conditions with masticatory performance and maximum bite force in older adults: A systematic review of cross-sectional studies. J Dent. 2022;123:104186.

34. Holmlund A, Lampa E, Lind L. Oral health and cardiovascular disease risk in a cohort of periodontitis patients. Atherosclerosis. 2017;262:101–106.

35. Joshipura KJ, Wand HC, Merchant AT, Rimm EB. Periodontal disease and biomarkers related to cardiovascular disease. J Dent Res. 2004;83:151–155.

36. Petrenya N, Hopstock LA, Holde GE, Oscarson N, Jönsson B. Relationship between periodontitis and risk of cardiovascular disease: insights from the tromsø study. J Periodontol. 2022;93:1353–1365.

37. Norhammar A, Näsman P, Buhlin K, de Faire U, Ferrannini G, Gustafsson A, Kjellström B, Kvist T, Jäghagen EL, Lindahl B, Nygren Å, Näslund U, Svenungsson E, Klinge B, Rydén L, PAROKRANK Study Group. Does Periodontitis Increase the Risk for Future Cardiovascular Events? Long-Term Follow-Up of the PAROKRANK Study. J Clin Periodontol. 2025;52:16–23.

38. Molina A, Martínez M, Montero E, Carasol M, Herrera D, Figuero E, Sanz M. Association between periodontitis and cardiovascular risk in spanish employed adults-the workers’ oral health study. J Periodontal Res. 2025;60:340–349.

39. J Y, L F, J R, G W, S C, Q Z, Z D, S Z, C H, X W, L L. Correlation between the severity of periodontitis and coronary artery stenosis in a chinese population. Aust Dent J. 2013;58. doi:10.1111/adj.12087.

40. Lee HJ, Choi EK, Park JB, Han KD, Oh S. Tooth loss predicts myocardial infarction, heart failure, stroke, and death. J Dent Res. 2019;98:164–170.

41. Pejcic A, Kostic M, Marko I, Obradovic R, Minic I, Bradic-Vasic M, Gligorijevic N, Kurtagic D. Tooth loss and periodontal status in patients with cardiovascular disease in the serbian population: a randomized prospective study. Int J Dent Hyg. 2023;21:317–327.

42. Matsuyama Y, Jürges H, Listl S. Causal Effect of Tooth Loss on Cardiovascular Diseases. J Dent Res. 2023;102:37–44.

43. Talekar DAW, Abraham DD, Puri DA, Gupta DA, Singal DK, Neha DN. Association between cardiovascular diseases and apical periodontitis: an umbrella review with stratification of evidence and sensitivity analysis. J Dent. 2025;:105943.

44. Chauhan N, Mittal S, Tewari S, Sen J, Laller K. Association of Apical Periodontitis with Cardiovascular Disease via Noninvasive Assessment of Endothelial Function and Subclinical Atherosclerosis. J Endod. 2019;45:681–690.

45. Ghanbari Z, Moradi Y, Samiee N, Moradpour F. Dental caries prevalence in relation to the cardiovascular diseases: cross-sectional findings from the iranian kurdish population. BMC Oral Health. 2024;24:509.

46. Ylöstalo PV, Järvelin MR, Laitinen J, Knuuttila ML. Gingivitis, dental caries and tooth loss: risk factors for cardiovascular diseases or indicators of elevated health risks. J Clin Periodontol. 2006;33:92–101.

47. Sen S, Logue L, Logue M, Otersen EAL, Mason E, Moss K, Curtis J, Hicklin D, Nichols C, Rosamond WD, Gottesman RF, Beck J. Dental Caries, Race and Incident Ischemic Stroke, Coronary Heart Disease, and Death. Stroke. 2024;55:40–49.

48. Sanchez P, Everett B, Salamonson Y, Ajwani S, Bhole S, Bishop J, Lintern K, Nolan S, Rajaratnam R, Redfern J, Sheehan M, Skarligos F, Spencer L, Srinivas R, George A. Oral health and cardiovascular care: Perceptions of people with cardiovascular disease. PLOS One. 2017;12:e0181189.

49. Liang L, Chen C-Y, Aris IM. Association of the Life’s Essential 8 cardiovascular health score with periodontitis among US adults. J Clin Periodontol. 2024;51:1502–1510.

50. Guo J-J, Hang Q-Q, Xu T, Liang W-X, Gao J-K, Ou H-B, Jiang F-Z, Qiu X-C-H, Tian Z-Z, Zhang Y-Z, Zhang J. Central adiposity indices and inflammatory markers mediate the association between life’s crucial 9 and periodontitis in US adults. Lipids Health Dis. 2025;24:199.

51. Ravera A, Carubelli V, Sciatti E, Bonadei I, Gorga E, Cani D, Vizzardi E, Metra M, Lombardi C. Nutrition and cardiovascular disease: finding the perfect recipe for cardiovascular health. Nutrients. 2016;8:363.

52. Tada A, Miura H. Systematic review of the association of mastication with food and nutrient intake in the independent elderly. Arch Gerontol Geriatr. 2014;59:497–505.

53. Karawekpanyawong R, Nohno K, Kubota Y, Ogawa H. Oral Health and Nutritional Intake in Community-Dwelling 90-Year-Old Japanese People: A Cross-Sectional Study. Gerodontology. 2023;40:100–111.

54. Altenhoevel A, Norman K, Smoliner C, Peroz I. The impact of self-perceived masticatory function on nutrition and gastrointestinal complaints in the elderly. J Nutr Health Aging. 2012;16:175–178.

55. Okamoto N, Amano N, Nakamura T, Yanagi M. Relationship between tooth loss, low masticatory ability, and nutritional indices in the elderly: a cross-sectional study. BMC Oral Health. 2019;19:110.

56. Schenkein HA, Loos BG. Inflammatory mechanisms linking periodontal diseases to cardiovascular diseases. J Clin Periodontol. 2013;40 Suppl 14:S51–69.

57. Dhungana G, Srisai D, Sampath C, Soliman J, Kelly RM, Saleh HY, Sedik A, Raynes E, Ferguson A, Alluri LSC, Gangula PR. Unveiling the molecular crosstalk between periodontal and cardiovascular diseases: a systematic review. Dent J. 2025;13:98.

58. Corredor Z, Suarez-Molina A, Fong C, Cifuentes-C L, Guauque-Olarte S. Presence of periodontal pathogenic bacteria in blood of patients with coronary artery disease. Sci Rep. 2022;12:1241.

59. Ziebolz D, Jahn C, Pegel J, Semper-Pinnecke E, Mausberg RF, Waldmann-Beushausen R, Schöndube FA, Danner BC. Periodontal bacteria DNA findings in human cardiac tissue - is there a link of periodontitis to heart valve disease? Int J Cardiol. 2018;251:74–79.

60. Aarabi G, Zeller T, Seedorf H, Reissmann DR, Heydecke G, Schaefer AS, Seedorf U. Genetic Susceptibility Contributing to Periodontal and Cardiovascular Disease. J Dent Res. 2017;96:610–617.

61. Kojima G, Taniguchi Y, Iwasaki M, Aoyama R, Urano T. Associations between self-reported masticatory dysfunction and frailty: a systematic review and meta-analysis. PLOS One. 2022;17:e0273812.

62. Okada M, Hama Y, Futatsuya R, Sasaki Y, Noritake K, Yamaguchi K, Matsuzaki M, Kubota C, Hosoda A, Minakuchi S. Association between masticatory performance, nutritional intake, and frailty in japanese older adults. Nutrients. 2023;15:5075.

63. Yamazaki T, Yamori M, Asai K, Nakano-Araki I, Yamaguchi A, Takahashi K, Sekine A, Matsuda F, Kosugi S, Nakayama T. Mastication and risk for diabetes in a japanese population: a cross-sectional study. PLOS One. 2013;8:e64113.

64. Hasegawa Y, Ono T, Sakagami J, Hori K, Maeda Y, Hamasaki T, Nokubi T. Influence of voluntary control of masticatory side and rhythm on cerebral hemodynamics. Clin Oral Investig. 2011;15:113–118.

65. Hasegawa Y, Ono T, Hori K, Nokubi T. Influence of human jaw movement on cerebral blood flow. J Dent Res. 2007;86:64–68.

66. Takeuchi N, Ekuni D, Tomofuji T, Morita M. Relationship between masticatory performance and heart rate variability: a pilot study. Acta Odontol Scand. 2013;71:807–812.

67. Farella M, Bakke M, Michelotti A, Marotta G, Martina R. Cardiovascular responses in humans to experimental chewing of gums of different consistencies. Arch Oral Biol. 1999;44:835–842.

68. Del Seppia C, Ghione S, Foresi P, Lapi D, Fommei E, Colantuoni A, Scuri R. Evidence in the human of a hypotensive and a bradycardic effect after mouth opening maintained for 10 min. Eur J Appl Physiol. 2017;117:1485–1491.

